# Reliability and Validity of the Japanese Version of the Survey of Perceived Organizational Support

**DOI:** 10.1101/2023.11.15.23298552

**Authors:** Kiminori Odagami, Tomohisa Nagata, Hisashi Eguchi, Akiomi Inoue, Kosuke Mafune, Koji Mori

**Affiliations:** Department of Occupational Health Practice and Management, Institute of Industrial Ecological Sciences, University of Occupational and Environmental Health, Japan, 1-1 Iseigaoka, Yahatanishi-ku, Kitakyushu 807-8555, Japan; Department of Mental Health, Institute of Industrial Ecological Sciences, University of Occupational and Environmental Health, Japan, 1-1 Iseigaoka, Yahatanishi-ku, Kitakyushu 807-8555, Japan; Institutional Research Center, University of Occupational and Environmental Health, Japan, 1-1 Iseigaoka, Yahatanishi-ku, Kitakyushu 807-8555, Japan

## Abstract

**Objectives:** This study aimed to validate the Japanese version of the Survey of Perceived Organizational Support (SPOS-J) for the Japanese workforce by examining its reliability, factorial, construct, and test-retest validity.

**Methods:** We conducted an online baseline survey with 6,220 employed Japanese individuals, followed by a follow-up survey two weeks later to examine the test-retest reliability of the SPOS-J. The SPOS-J was translated in accordance with the ISPOR Task Force Guidelines. Both confirmatory and exploratory factor analyses were utilized to examine factorial validity. Construct validity was assessed using Pearson’s correlation analysis, while internal consistency was determined through the calculation of Cronbach’s alpha values. Additionally, test-retest reliability was evaluated with Cohen’s weighted kappa and intraclass correlation coefficients.

**Results:** The SPOS-J demonstrated a better fit for a two-factor structure, comprising two subscales - SPOS-J (supported) and SPOS-J (unsupported) – compared to the one-factor structure of the original version. It exhibited high internal consistency, with Cronbach’s alpha coefficients of .93 for the overall SPOS-J, .97 for the SPOS-J (supported), and .95 for the SPOS-J (unsupported). Test-retest reliability was found to be good for the overall SPOS-J (.62) and moderate for its subscales, with Cohen’s weighted kappa values of .54 for SPOS-J (supported) and .42 for SPOS-J (unsupported). The construct validity was confirmed with significant correlations between the SPOS-J, its antecedents, and outcomes.

**Conclusions:** The SPOS-J is a reliable and valid measure for assessing perceived organizational support among Japanese workers.

## 1. INTRODUCTION

Perceived Organizational Support (POS) is a concept proposed by Eisenberger et al. ^1^ in 1986, and is defined as "global beliefs concerning the extent to which the organization values their contributions and cares about their well-being." A major characteristic of POS is that it focuses not on the company’s (management and human resource management department’s) point of view, but on the employee’s point of view, i.e., how the employees who work there feel about the company. POS is based on social exchange theory, the theory that human beings form and develop relationships through the exchange of various tangible and intangible rewards^1^. Therefore, it is based on the idea that the more support an employee feels from the organization, the more favorable the employee feels toward the organization, and the more the employee will make further efforts to contribute to the achievement of the organization’s goals. Therefore, POS is seen as a concept with important benefits for both employees and organizations, and research on POS began to increase in the mid-1990s and has progressed rapidly in recent years.

Studies have shown a relationship between POS and a variety of health-related indicators. Antecedents of POS include procedural justice^2^ (i.e., one of the sub-concepts of organizational justice), supervisor support^3,4^, organizational rewards^5^, and job conditions^6–8^, while outcome factors include emotional commitment to the organization^3^, job satisfaction^9^, performance^10,11^, and withdrawal behavior^4,12^. Many of these antecedent and outcome factors are positively correlated with POS. On the other hand, antecedent factors such as job conditions (role ambiguity^7^, role conflict^8^, size of organization^7^) and organizational politics^13^, and outcome factors such as withdrawal behavior (turnover intentions^12^, turnover^4^) and strains are negatively correlated with POS.

A representative measure of POS is the Survey of Perceived Organizational Support (SPOS)^14^. The SPOS is a 36-question measure developed by Eisenberger et al. in 1986^1^. In much of the past literature, the original version of the SPOS was one-dimensional and had high internal reliability, leading to the use of shortened versions such as the 16-item version^1^ and the 8-item version^6^. Studies using the SPOS have been conducted mainly in English-speaking countries, European countries, and China, and besides the English version, Malay^15^ and Turkish^16^ versions of the SPOS have been developed. However, there is currently no Japanese version of the SPOS that has been tested for reliability and validity; therefore, studies have been conducted using original questionnaire items that reflect the POS concept^17^, and it has not been possible to compare findings from Japanese studies with those from foreign studies.

Currently, in many developed countries, including Japan, the working-age population is declining due to the low birthrate and aging population^18^. Therefore, companies are faced with the need to make strategic efforts to improve POS by showing active consideration for the well-being of their employees, and to aim at improving corporate performance and curbing employee turnover behavior through improving employee job satisfaction and work engagement. Furthermore, since companies need to build good relationships with diverse employees due to the recent diversification of employee employment patterns and needs, concepts, and indicators that reflect employees’ evaluations and opinions of the company, such as POS, are considered useful in verifying the effectiveness of personnel policies that aim to absorb diversity.

The purpose of this study is to examine the reliability and validity of the Japanese version of the SPOS (SPOS-J) for the general workforce in Japan. More specifically, we examined its factorial and construct validity as well as its reliability (i.e., internal consistency and test-retest reliability).

## 2. METHODS

### 2.1 Participants

A baseline online survey was conducted from January 24 to February 3, 2022, among registrants of Cross Marketing Inc. (Tokyo, Japan), a private online survey firm in Japan. As of January 2022, there were approximately 2.95 million registered participants, from which 102,032 were randomly selected to receive participation invitations via e-mail. Of these, 15,676 responded to the preliminary survey and participated in the main survey (baseline survey) if they met all of the screening criteria: (1) age 20–64, (2) currently working, and (3) employed (full-time, contract, temporary, part-time, or casual employee).

For this baseline survey, the sample was collected based on data from the 2020 Labor Force Survey (Ministry of Internal Affairs and Communications), stratified by gender (men: 54.4%, women: 45.6%), age (20–29: 18.6%, 30–39: 21.1%, 40–49: 27.7%, 50–59: 24.0%, 60–64: 8.7%), and region of residence (Hokkaido: 3.8%, Tohoku: 6.5%, South Kanto: 31.6%, North Kanto/Koshin: 7.4%, Hokuriku: 4.1%, Tokai: 12.2%, Kinki: 15.7%, Chugoku: 5.5%, Shikoku: 2.6%, Kyushu/Okinawa: 10.5%). Responses to this survey were on a first-come, first-served basis, and due to budget constraints, the target number of respondents was set at 6,000, and once the final 6,220 respondents were reached, the recruitment process was closed because the sample was collected at a stratified percentage.

In addition, to examine the test-retest reliability of the SPOS-J, a follow-up study using the SPOS-J was conducted two weeks after the baseline study. The follow-up survey was administered to those respondents to the baseline survey who answered "no" to both (1) "changed workplaces within the past two weeks" and (2) "had a significant change in workplaces within the past two weeks" at the time of their follow-up survey responses. As in the baseline survey, responses were on a first-come, first-served basis, and the survey was closed when the number of respondents reached 452. Because the online survey required all questions to be answered, no participants had incomplete responses.

### 2.2 Measures

#### 2.2.1 SPOS-J

The Japanese version of SPOS was prepared in accordance with International Society for Pharmacoeconomics and Outcomes Research (ISPOR) Task Force Guidelines^19^. First, permission for Japanese translation was obtained from the original author (Robert Eisenberger), and the original version of SPOS^1^ was independently translated into Japanese by four native Japanese-speaking researchers (KO, TN, HE, and KM), and a draft Japanese version was prepared after discussion among the researchers. Next, native experts who had not read the original text were asked to translate the draft Japanese version into English. The original author reviewed the back-translated English version, and the draft Japanese version was revised. Since there is no corresponding Japanese word for "well-being" and the specific image of "well-being" varies from person to person, we consulted with the original author and added the definition from the Longman Dictionary of Contemporary English^20^, adding that it is in the context of work, "a feeling of being comfortable, healthy, and happy through work" was added as a supplementary explanation. Next, as a cognitive debriefing, we asked five workers of various occupations and ages collected through snowball sampling for their opinions by asking them, "Please advise us if you have any difficulty understanding or answering any questions in Japanese. As a result, minor comments were received, including suggestions for the use of more easily understood verbs and adjectives. The four researchers considered the pros and cons of reflecting these comments in the translation, and adjusted the translation so that it would not affect the meaning when translated into English. The Japanese version of SPOS (SPOS-J) was thus completed.

The SPOS-J consists of 36 items, and as in the original version of the SPOS, respondents were asked to respond to each item on a 7-point scale (0 = strongly disagree to 6 = strongly agree). Of the 36 items, 18 items asked for positive content, while the remaining 18 items asked for negative content. The scoring of the latter items asking about negative content was reversed, with higher scores for all items indicating a higher POS. Finally, the average score for the 36 items was calculated.

#### 2.2.2 Relevant Variables

In this study, the following indicators were selected based on the fact that existing studies have shown that these indicators have a strong correlation with POS^14^ and that valid Japanese language scales are available for use. Role conflict from job conditions and intention to leave from withdrawal behavior were selected as indicators of negative correlation with POS.

##### Possible antecedents

・Supervisor Support : The corresponding subscale of the New Brief Job Stress Questionnaire (New BJSQ) was used. The New BJSQ has been validated with Japanese workers and found to have acceptable levels of internal consistency, reliability, and construct validity on almost all subscales^21^. Three items from the New BJSQ regarding supervisor support (e.g., How freely can you talk with your supervisor?) were rated using a 4-point scale (1 = extremely to 4 = not at all). The scores were reversed in the analysis to make them more intuitive and easier to understand, and the average score for the three items was calculated. The Cronbach’s alpha coefficient for this scale was .88.

・Organizational Justice (procedural justice/ interactional justice) : Using the Japanese version of the Organizational Justice Scale^22^, which is the Japanese version of the modified version of the Organizational Justice Scale^23^ developed by Moorman^24^, we rated each of the two subscales, procedural justice (7 items) and interactional justice (6 items), using a 5-point scale (1 = strongly disagree to 5 = strongly agree) and calculated the mean score. Cronbach’s alpha coefficients for these subscales were .94 and .96 for procedural and interaction justice, respectively.

・Role conflict : One item on the New BJSQ ("I receive incompatible requests from two or more people") was rated using the four-point scale (1 = definitely to 4 = not at all), and the scores were reversed in the analysis for easier understanding.

##### Possible outcomes

・Job Satisfaction : One item ("I am satisfied with my job") from the New BJSQ was used to evaluate the results. This indicator was rated using a 4-point scale (1 = satisfied to 4 = dissatisfied), and the scores were reversed in the analysis for ease of understanding.

・Organizational (Affective) commitment : Three items from the Japan Institute of Labour (Current The Japan Institute for Labour Policy and Training) Human Resource Management checklist^25^ regarding affective commitment (e.g., "I feel as if this company’s problems are my own problems") were evaluated using a 5-point scale (1 = not at all true to 5 = very true), and the average score was calculated. The Cronbach’s alpha coefficient for this scale was .73.

・Turnover intention : A one-question shortened version of the Japanese version of the Turnover Intention Scale^26^ ("How often have you seriously considered quitting your current job?") was used and rated using a 6-point scale (1 = not at all to 6 = very often).

#### 2.2.3 Demographic Characteristics

Participant demographic items were collected at the time of the baseline survey. In addition to gender (men, women) and age, respondents were asked to provide their final education (junior high school/high school, vocational school/junior college, university/graduate school), household income (< 3.00 million Japanese yen (JPY), 3.00–4.99 million JPY, 5.00–9.99 million JPY, and 10.00 million JPY or greater), and job type (administrative, professional/technical, clerical, sales, service, security, agriculture/forestry/fishing, production process jobs, transportation/machine operation, construction/mining, transportation/cleaning/packaging, and others) based on the Japan Standard Occupational Classification (statistical standard established in December 2009).

### 2.3 Statistical Analysis

A confirmatory factor analysis was performed using a one-factor model for SPOS-J based on the original version of SPOS^1^ by Eisenberger et al. Model fit was evaluated using a combination of goodness-of-fit indices, including χ^2^ goodness-of-fit statistic, Root Mean Square Error of Approximation (RMSEA), Akaike’s Information Criterion (AIC), Comparative Fit Index (CFI), and Standardized root mean squared residual (SRMR). Acceptable model fit criteria were defined as RMSEA and SRMR < 0.08 and CFI > 0.90^27^. In case the results of the confirmatory factor analysis showed a poor fit of the one-factor model, an exploratory factor analysis was conducted. Exploratory factor analysis (maximum likelihood method, promax rotation) was performed on all 36 items of the SPOS-J, and the number of factors was determined at the junction where the cumulative contribution exceeds 50%. A confirmatory factor analysis was performed on the factor structure supported by the exploratory factor analysis, and the model goodness of fit was compared to the one-factor structure of the original version.

To assess construct validity, Pearson’s correlation coefficients were calculated for the SPOS-J (including subscales) with the antecedents (supervisor support, organizational justice, and role conflict) and outcome factors assumed above (job satisfaction, organizational commitment, and turnover intention). Following Cohen^28^, we describe effects as small (0.10), medium (0.30), and large (0.50). To assess internal consistency, we calculated Cronbach’s alpha coefficients for the SPOS-J and its subscales. To assess test-retest reliability, the Kolmogorov-Smirnov Test was first used to check whether the SPOS-J and its subscales, each of the 36 items, followed a normal distribution. Since these were found not to follow a normal distribution, Cohen’s linearly weighted kappa coefficients were calculated for each and evaluated using Landis & Koch criteria^29^(poor: -1.00 to 0.20; reasonable: 0.20 to 0.40; moderate: 0.41 to 0.60; good: 0.61 to 0.80; and very good: 0.81 to 1.00). The intraclass correlation coefficient (ICC [1,1]) and standard error of measurement (SEM) were calculated as parametric statistics for test-retest reliability as reference values. Data from 452 individuals who responded to the follow-up survey were used to calculate Cohen’s weighted kappa coefficient, ICC, and SEM.

The significance level was set at 0.05 (two-tailed). All analyses were performed using IBM SPSS Statistics Version 27.0, Stata version 16.

## 3. RESULTS

### 3.1 Characteristics of participants

Table 1 shows the number of participants in the baseline and follow-up surveys and the characteristics of the participants. In the baseline survey, participants had the same gender and age distribution as in the data from the 2020 Labor Force Survey (Ministry of Internal Affairs and Communications), and participants’ annual household income of 5.00–9.99 million JPY was the highest (43.2%), and 50.5% of participants had a college degree or higher as their final education level. The most common occupations were clerical, professional/technical, and service, in that order. There were no significant differences in gender, age, final education, or occupation between baseline and follow-up.

**Table 1.** Demographics of participants at 1st survey and 2nd survey.

| Demographic Characteristics | Baseline |  | Follow-Up<sup>※</sup> |  |
| --- | --- | --- | --- | --- |
|  | n (%) |  | n (%) |  |
| Total | 6220 |  | 452 |  |
| Gender |  |  |  |  |
| Men | 3382 | (54.3) | 248 | (54.9) |
| Women | 2838 | (45.6) | 204 | (45.1) |
| Age |  |  |  |  |
| 20-29 | 1131 | (18.2) | 59 | (13.1) |
| 30-39 | 1281 | (20.6) | 66 | (14.6) |
| 40-49 | 1684 | (27.1) | 142 | (31.4) |
| 50-59 | 1455 | (23.4) | 108 | (23.9) |
| 60-64 | 669 | (10.8) | 77 | (17.0) |
| Annual household income (million yen) |  |  |  |  |
| <3.00 | 1073 | (17.3) | 74 | (16.4) |
| 3.00-4.99 | 1658 | (26.7) | 128 | (28.3) |
| 5.00-9.99 | 2689 | (43.2) | 185 | (40.9) |
| ≥10.00 | 800 | (12.9) | 65 | (14.4) |
| Final Education |  |  |  |  |
| Junior high or high school | 1748 | (28.1) | 128 | (28.3) |
| Vocational school or college | 1331 | (21.4) | 91 | (20.1) |
| University or graduate school | 3141 | (50.5) | 233 | (51.6) |
| Job type |  |  |  |  |
| Administrative | 402 | (6.5) | 35 | (7.7) |
| Professional and technical | 1095 | (17.6) | 67 | (14.8) |
| Clerical | 1702 | (27.4) | 123 | (27.2) |
| Sales | 616 | (9.9) | 46 | (10.2) |
| Service | 842 | (13.5) | 54 | (12.0) |
| Security | 91 | (1.5) | 7 | (1.6) |
| Agriculture, forestry, and fishing | 16 | (0.3) | 3 | (0.7) |
| Production process jobs | 459 | (7.4) | 42 | (9.3) |
| Transportation and machine operation | 115 | (1.9) | 8 | (1.8) |
| Construction and mining | 91 | (1.5) | 4 | (0.9) |
| Transportation, Cleaning, Packaging | 217 | (3.5) | 19 | (4.2) |
| Others | 574 | (9.2) | 44 | (9.7) |

※ Follow-up sample was used only to examine the test-retest reliability.

**Table 2.** Items, means, standard deviations, and exploratory factor analysis with maximum likelihood method and promax rotation (N=6,220)

| Variable | Mean | SD | Factor Loadings |  | Communality |
| --- | --- | --- | --- | --- | --- |
|  |  |  | 1 | 2 |  |
| Item_25 | 2.88 | 1.35 | 0.87 | 0.06 | 0.75 |
| Item_10 | 2.68 | 1.34 | 0.87 | 0.05 | 0.75 |
| Item_9 | 2.62 | 1.34 | 0.86 | 0.06 | 0.73 |
| Item_21 | 2.63 | 1.34 | 0.85 | 0.05 | 0.73 |
| Item_20 | 2.84 | 1.31 | 0.82 | 0.00 | 0.68 |
| Item_27 | 2.71 | 1.28 | 0.82 | 0.00 | 0.68 |
| Item_36 | 2.67 | 1.33 | 0.82 | 0.05 | 0.67 |
| Item_8 | 2.81 | 1.38 | 0.82 | 0.02 | 0.67 |
| Item_4 | 2.67 | 1.37 | 0.81 | 0.04 | 0.65 |
| Item_1 | 2.69 | 1.46 | 0.79 | 0.08 | 0.63 |
| Item_33 | 2.76 | 1.28 | 0.79 | -0.04 | 0.63 |
| Item_35 | 2.39 | 1.31 | 0.73 | 0.02 | 0.53 |
| Item_13 | 3.06 | 1.38 | 0.72 | -0.09 | 0.53 |
| Item_18 | 2.57 | 1.30 | 0.72 | -0.05 | 0.52 |
| Item_29 | 2.84 | 1.31 | 0.70 | -0.09 | 0.51 |
| Item_5 | 3.02 | 1.51 | 0.68 | -0.04 | 0.46 |
| Item_30 | 2.38 | 1.41 | 0.68 | 0.03 | 0.46 |
| Item_24 | 2.73 | 1.42 | 0.67 | -0.01 | 0.45 |
| Item_17 | 3.11 | 1.45 | 0.12 | 0.82 | 0.68 |
| Item_23 | 3.21 | 1.39 | 0.09 | 0.81 | 0.66 |
| Item_7 | 3.28 | 1.34 | 0.09 | 0.79 | 0.62 |
| Item_6 | 3.43 | 1.41 | 0.15 | 0.79 | 0.63 |
| Item_32 | 3.07 | 1.41 | 0.05 | 0.76 | 0.58 |
| Item_3 | 3.18 | 1.46 | 0.08 | 0.76 | 0.58 |
| Item_15 | 3.48 | 1.29 | -0.01 | 0.74 | 0.54 |
| Item_14 | 3.45 | 1.33 | -0.04 | 0.72 | 0.52 |
| Item_12 | 3.27 | 1.37 | -0.09 | 0.70 | 0.51 |
| Item_19 | 2.83 | 1.48 | -0.06 | 0.69 | 0.48 |
| Item_16 | 2.77 | 1.65 | -0.01 | 0.68 | 0.46 |
| Item_22 | 3.65 | 1.39 | 0.02 | 0.67 | 0.45 |
| Item_28 | 2.94 | 1.44 | -0.10 | 0.67 | 0.47 |
| Item_34 | 3.35 | 1.44 | -0.04 | 0.65 | 0.43 |
| Item_2 | 3.11 | 1.53 | -0.08 | 0.64 | 0.42 |
| Item_31 | 3.10 | 1.40 | -0.15 | 0.61 | 0.40 |
| Item_26 | 3.82 | 1.34 | 0.00 | 0.59 | 0.35 |
| Item_11 | 3.49 | 1.44 | -0.03 | 0.59 | 0.35 |
| Inter-factor correlations |  | 1 | 1 |  |  |
|  |  | 2 | -0.04 | 1 |  |
| Eigenvalues |  |  | 11.11 | 9.07 |  |

### 3.2 Factorial validity

Based on the original version of the SPOS, a confirmatory factor analysis was performed on the SPOS-J with a one-factor model, and the model fit was significantly lower (χ^2^ = 85197; P < 0.001, RMSEA = 0.151, SRMR = 0.245, CFI = 0.511, AIC = 85341), so an exploratory factor analysis was performed. The number of factors was determined at the boundary point where the cumulative contribution exceeds 50%, and the number of factors was set to 2. The results of exploratory factor analysis (maximum likelihood method, promax rotation) under the assumption of two factors are shown (Table 2). The results indicate that the SPOS-J has a two-factor structure, with the first factor being items asking for supportive content (e.g., my organization values my opinion) and the second factor being items asking for non-supportive content (e.g., my organization shows little interest in my interests). Therefore, the first factor was designated SPOS-J (supported), and the second factor was designated SPOS-J (unsupported). A confirmatory factor analysis was performed on the 2-factor model, and the model fit was higher than the 1-factor (χ^2^ = 24706; P < 0.001, RMSEA = 0.081, SRMR = 0.067, CFI = 0.861, AIC = 24852), but not sufficient in terms of the acceptable model fit criteria set in this study (RMSEA, SRMR < 0.08, CFI > 0.90) (Table 3).

**Table 3.** Results of confirmatory factor analyses: Comparison of goodness-of-fit indices among one-factor and two-factor models (N=6,220)

| | $\chi^2$ | df | P | RMSEA (90%CI) | SRMR | CFI | AIC |
| --- | --- | --- | --- | --- | --- | --- | --- |
| One-factor model | 85197 | 594 | <.001 | 0.151 (0.150 - 0.152) | 0.245 | 0.511 | 85341 |
| Two-factor model | 24706 | 593 | <.001 | 0.081 (0.080 - 0.082) | 0.067 | 0.861 | 24852 |

RMSEA: root mean square error of approximation; SRMR: standardized root mean square residual; CFI: comparative fit index; AIC: Akaike's Information Criterion

### 3.3 Construct validity

Table 4 shows the correlations between the SPOS-J and its two subscales, SPOS-J (supported) and SPOS-J (unsupported), and all study variables, including antecedents (supervisor support, organizational justice [procedural and interactional], and role conflict) and outcome factors (job satisfaction, organizational commitment, and turnover intention). The two subscales showed a small negative correlation with role conflict, a small to medium negative correlation with turnover intention, and small to high positive correlations with other antecedents and outcome factors.

**Table 4.** Means, standard deviations and correlations of variables used in the study (n=6,220)

|  | Mean | SD | 1 | 1-1 | 1-2 |
| --- | --- | --- | --- | --- | --- |
| 1. SPOS-J | 2.99 | 0.73 |  |  |  |
| 1-1 SPOS-J ( supported ) | 2.72 | 1.07 | .71** |  |  |
| 1-2 SPOS-J ( unsupported ) | 3.25 | 1.03 | .68** | -.03* |  |
| 2. Supervisor Support | 2.68 | 0.81 | .52** | .45** | .27** |
| 3. Organizational justice ( procedural ) | 3.13 | 0.81 | .57** | .45** | .33** |
| 4. Organizational justice ( interactional/relational ) | 3.23 | 0.91 | .59** | .50** | .32* |
| 5. Role Conflict | 2.34 | 0.85 | -.23** | -.21** | -.12* |
| 6. Job Satisfaction | 2.54 | 0.83 | .46** | .40** | .24* |
| 7. Organizational Commitment | 2.80 | 0.83 | .43** | .46** | .13** |
| 8. Turnover intention | 2.69 | 1.38 | -.39** | -.35** | -.20** |

Note; \*p<.05, \*\*p<.01

### 3.4 Reliability (internal consistency and test-retest reliability)

Table 5 shows the Cronbach’s alpha coefficients, Cohen’s weighted kappa coefficients, ICCs, and SEM for SPOS-J, SPOS-J (supported), and SPOS-J (unsupported). Cronbach’s alpha coefficients for SPOS-J, SPOS-J (supported), and SPOS-J (unsupported) showed high consistency (.93, .97, and .95, respectively). In addition, Cohen’s weighted kappa coefficients and ICCs showed good test-retest reliability for SPOS-J and moderate test-retest reliability for SPOS-J (supported) and SPOS-J (unsupported) in the 2-week interval test-retest.

**Table 5.** Means, standard deviations, Minimum, Maximum, Cronbach’s α coefficients, Cohen’s weighted κ coefficients intraclass correlation coefficients, and standard error of measurement for the SPOS-J and its subscales

| | Mean | SD | Min | Max | Cron-<br>bach's $\alpha$ | Cohen's<br>Weighted $\kappa$<br>(95% CI) | ICC<br>(95% CI) | SEM<br>(95% CI) |
| --- | --- | --- | --- | --- | --- | --- | --- | --- |
| SPOS-J | 2.99 | 0.73 | 0 | 6 | 0.93 | 0.62<br>(0.58-0.67) | 0.81<br>(0.77 - 0.84) | 0.44<br>(0.41-0.48) |
| SPOS-J<br>(supported) | 2.72 | 1.07 | 0 | 6 | 0.97 | 0.54<br>(0.49-0.59) | 0.73<br>(0.68 - 0.77) | 0.68<br>(0.62-0.73) |
| SPOS-J<br>(unsupported) | 3.25 | 1.03 | 0 | 6 | 0.95 | 0.42<br>(0.36-0.48) | 0.51<br>(0.44 - 0.57) | 0.84<br>(0.78-0.90) |

ICC: intraclass correlation coefficients, SEM: standard error of measurement
CI: Confidence interval.
Note; Mean, SD, Min, Max, and Cronbach's $\alpha$ were calculated using all responses in the baseline survey.

## 4. DISCUSSION

### 4.1 Reliability and validity

This study created the SPOS-J and verified its reliability and validity. To assess factorial validity, confirmatory and exploratory factor analyses were conducted, and to assess construct validity, the SPOS-J was examined in relation to antecedent and outcome factors identified from existing research. Furthermore, internal consistency and test-retest reliability were examined to assess the reliability of the SPOS-J.

Regarding Factorial validity, the two-factor structure consisting of two subscales, SPOS-J (supported) and SPOS-J (unsupported), was a better fit than the one-factor structure in the original version. SPOS-J (supported) items asked for supportive content, while SPOS-J (unsupported) items asked for non-supportive content. This result may be related to the cultural background of the Japanese. While the West has an independent construal of self, Japan is known to have a predominantly interdependent construal of self, and a culture shaped by an interdependent construal of self will tend toward self-critical self-evaluation, focusing on its own weaknesses and problems in comparison to the ideal role image by society^30^. In fact, it has been reported that in Japan, there is a self-critical tendency to evaluate oneself lower or more negatively than others^31^. This tendency toward self-criticism may be the result of low self-estimation as well as high estimation of others, and Endo^32^ reported the possibility that Japanese people are simultaneously lowering their self-estimation and raising their evaluation of others. In the SPOS-J survey, respondents were asked to evaluate their own organizations, and from their perspective, the organizations to which they belonged could be considered as others. Therefore, it is possible that respondents gave stronger positive evaluations of their own organizations and weaker negative evaluations of others, resulting in a two-factor structure that eliminates the one-dimensionality of the results.

In this validation, the model fit was not necessarily sufficient even for the two-factor model. One reason for this is that the present validation was conducted using all 36 items of the SPOS-J. These items may include questions that are inappropriate or meaningless depending on the country’s culture, employment system, or specific industries (e.g., civil servants and other industries with high employment security). In fact, when examining the reliability and validity of the SPOS for teachers, Dolma et al.^16^ excluded items from the 36 items of the SPOS that were not appropriate for their employment environment as teachers. The original version of the SPOS^1^ by Eisenberger et al. was basically developed to understand POS for U.S. workers. To develop POS research in Japan in the future, it is necessary to improve the scale to make it more suitable for Japanese workers by removing items that do not fit Japanese culture and labor practices.

Regarding construct validity, this study examined the relationship between supervisor support, organizational justice [procedural and interactional], and role conflict as antecedents of the SPOS-J. The SPOS-J was highly correlated with supervisor support (r = .52). Several studies examining supervisor support as a variable affecting POS have confirmed a highly positive relationship between supervisor support and POS^3,4,33^, which is consistent with the results of this study. Regarding organizational justice, this study confirmed the relationship between the SPOS-J and procedural and interactional justice. We found a high positive correlation (r = .57) between procedural justice and SPOS-J, which is consistent with the finding of Moorman et al^2^. Interactional justice has also been reported to be positively correlated with POS^34^. The results of a meta-analysis by Rhoades & Eisenberger ^14^ also reported a high correlation between interactional justice and POS, consistent with the results of this study (r = .59). Stressors are another antecedent of POS, but research on POS has focused primarily on the effects of stressors related to their role in the organization. Regarding role conflict, a role-related stressor, several studies^8,35^ have shown that role conflict and POS are negatively correlated, but the strength of the correlation ranges (r = -.20 to -.64). The results of this study showed a medium negative correlation (r = -.23), which was close to the results of Hutchison et al.^35^.

We examined the relationship between job satisfaction, organizational (affective) commitment, and turnover intention as outcome factors on the SPOS-J. Job Satisfaction has been empirically found to have a medium positive correlation with POS^9^ and was generally consistent with the results of this study (r = .46). Based on social exchange theory and the idea of reciprocity norms, Eisenberger et al. ^1^ argue that POS can foster a sense of obligation to the organization and increase affective commitment to the organization. The results of this study also showed that POS had a middle positive correlation (r = .43) with affective commitment, and many existing studies^5,10,36^ found a positive relationship between POS and affective commitment, although the strength of the correlation varied(r = .38 to .81). This variation may have been caused by differences in the target population. In a meta-analysis by Rhoades & Eisenberger ^14^, among withdrawal behaviors (turnover intentions, turnover, and other withdrawal behaviors), the strongest relationship with POS was found for turnover intentions, and turnover intentions have been empirically shown to have a middle negative correlation with POS^12^. This result was consistent with the results of this study (r = -.39).

The SPOS-J (unsupported), a subscale of the SPOS-J, had weaker correlations with all antecedent and outcome factors than the SPOS-J or another subscale, the SPOS-J (supported). This result may be due to the tendency of Japanese people to evaluate negative evaluations of other organizations more weakly because of their cultural background^30,31^.

For reliability, the internal consistency of SPOS-J, SPOS-J (supported), and SPOS-J (unsupported) was sufficient with Cronbach’s alpha coefficients (.94, .92, and .87, respectively), and SPOS-J (36 items) was equivalent to the original version (36 items)^1^. Test-retest reliability was confirmed to be good for SPOS-J and moderate for SPOS-J (supported) and SPOS-J (unsupported), as indicated by Cohen’s weighted kappa coefficient and ICC. To the best of our knowledge, we could not find any studies that examined test-retest reliability abroad, but this study suggests that, at least for the SPOS-J, it has a certain level of test-retest reliability.

### 4.2 Practical Implications

In recent years, there has been a great deal of research on the relationship between work engagement, which has received attention as a predictor of physical and mental health^37^ and job performance^38^, and POS, and work engagement has been found to be positively related to POS^39^. The concept of strategically working to improve POS by showing active consideration for employee well-being, and aiming to improve corporate performance and curb employee turnover by improving employee job satisfaction and work engagement, is highly compatible with the concept of health and productivity management^40^ that is gaining popularity in Japan. Therefore, making SPOS-J available as an evaluation indicator for organizations may contribute to the further development of occupational health practice and health and productivity management in Japan though the provision of a new evaluation indicator.

### 4.3 Limitations

This study has several limitations. First, this study is an Internet-based survey and may not represent the general population of Japanese workers, since those who do not have Internet access or are not registered with an online survey firm are omitted from the target population. However, since this study is based on data from a labor force survey conducted by a public agency, and the sample was collected stratified by gender, age, and region of residence to reduce sampling bias, the generalizability of the study’s results is considered sufficient. Second, this study is based on survey data from self-reports, and the present results should be replicated in the future using objective measures (e.g., measures related to performance, leaving a job, and other behaviors). Third, this study did not assess known-groups validity, criterion validity, cross-cultural validity, or responsiveness.

## 5. CONCLISION

In conclusion, this study confirms that the Japanese version of SPOS (SPOS-J) can adequately measure POS and can be used in a Japanese context.

## ACKNOWLEDGEMENTS

This study was initiated with the permission of the original author, Dr. Robert Eisenberger, for the preparation of SPOS-J and was completed after his review. He subsequently passed away and we were unable to obtain confirmation of this paper. We thank Dr. Robert Eisenberger for his expert assistance as the original author in the process of developing the Japanese version of the scale.

## AUTHOR CONTRIBUTIONS

KO and TN, HE, KosM, KojM designed the study; KO and TN, HE, KojM participated in the development of the Japanese version of the scale; KO and KojM, AI analyzed the data; KO led the writing of the manuscript, and all other authors were involved in the drafting the manuscript. All authors read and approved the final manuscript.

## DATA AVAILABILITY STATEMENT

The data that support the findings of this study are available from the corresponding author upon reasonable request.

